# Towards the elimination of mother-to-child transmission of syphilis in the WHO European region: Implications from a survey in the non-European Union and European Economic Area countries

**DOI:** 10.1101/2025.04.09.25325528

**Authors:** Machiko Otani, Jane Rowley, Giorgi Kuchukhidze, Viatcheslav Grankov, Stela Bivol, the WHO European Region non-EU/EEA STI Surveillance network

## Abstract

Globally, the public health burden of congenital syphilis (CS) remains high despite efforts to eliminate mother-to-child transmission (EMTCT) of syphilis and the fact that this disease is treatable if diagnosed early. In the WHO European region, the burden is not fully available, as CS cases are not systematically reported by the non-EU/EEA countries, while data from the European Union and European Economic Area (EU/EEA) countries are regularly reported by the European Centre for Disease Prevention and Control. In 2023, only 8 out of 24 non-EU/EEA countries reported the number of CS cases to Global AIDS Monitoring. In 2024, the WHO Regional Office for Europe conducted a survey among the 24 non-EU/EEA countries. Based on the data from 19 countries, the total number of CS cases was 93 (2.8 per 100,000 live births) in 2023. Seven countries reported no cases between 2021 and 2023, and four countries accounted for 84% of the cases. The actual burden of CS could be higher because of the suboptimal surveillance systems and variation in case definition used for surveillance. The survey results underline the importance of strengthening CS surveillance in non-EU/EEA countries, including standardising the case definition and improving completeness and coverage.

## Background

In 2007, the World Health Organization (WHO) and its partners launched the global initiative for the elimination of mother-to-child transmission (EMTCT) of syphilis and human immunodeficiency virus (HIV). In 2022, this initiative expanded to include hepatitis B and became a “triple elimination” initiative (1). As of February 2025, 18 countries and territories have been validated for EMTCT of syphilis(2). The validation criteria include achieving a congenital syphilis (CS) case rate of ≤50 per 100,000 live births and reaching three programmatic targets: antenatal care (ANC) coverage (defined as at least one visit) ≥95%, syphilis testing coverage among pregnant women attending ANC ≥95%, and syphilis treatment coverage among syphilis-seropositive pregnant women ≥95% (3).

The impact indicator for EMTCT is based on the global surveillance case definition of CS (3) (see Box 1).

### Box 1. Congenital syphilis case definitions

| Global surveillance case definition | 2018 EU case definition |
| --- | --- |
| *\* Depending on the clinical context, either of the two following options for the surveillance case definition of CS may be used*<br>[1] a live birth or foetal death at >20 weeks of gestation or >500 g (including stillbirth) born to a woman with positive syphilis serology and without adequate syphilis treatment<br>**OR**<br>[2] a live birth, stillbirth or child <2 years of age born to a woman with positive syphilis serology or with unknown serostatus and with laboratory and/or radiographic and/or clinical evidence of syphilis infection (regardless of the timing or adequacy of maternal treatment) | *\* Only cases in infants that meet the laboratory criteria for case confirmation are currently under EU epidemiological surveillance*<br>**Criteria for laboratory confirmation**<br>At least one of the following three:<br>— Demonstration of *Treponema pallidum* by dark field microscopy in the umbilical cord, the placenta, a nasal discharge or skin lesion material<br>— Demonstration of *Treponema pallidum* by DFA-TP in the umbilical cord, the placenta, a nasal discharge or skin lesion material<br>— Detection of *Treponema pallidum*-specific IgM (FTA-abs, EIA)<br>AND a reactive non-treponemal test (VDRL, RPR) in the child's serum |

According to the WHO estimation, the WHO European Region has the lowest CS case rate of the six WHO regions(4), 3.5 per 100,000 live births in 2022 (unpublished data), and is the only region with a CS case rate of less than 50 per 100,000 live births. WHO European Regional Action Plan for HIV, viral hepatitis and STIs has an ambitious target of reducing this further to <= 1.0 case per 100,000 live births in 2030 (5). As of February 2025, two countries in the WHO European region, Belarus and the Republic of Moldova, have been validated for EMTCT of syphilis and have maintained their status (2).

The European Centre for Disease Prevention and Control (ECDC) collects data and publishes an epidemiological report on reported CS cases annually that covers 30 of the WHO European member states. According to their latest report, which draws on data from 26 countries, there were 78 CS cases in 2023 or a case rate of 2.7 per 100,000 live births(6). For the 24 non-EU/EEA countries in the WHO European region, annual data collection has been through the Global AIDS Monitoring (GAM) (7). Eleven out of the 24 countries provided data on the number of reported CS cases at least once in either 2021, 2022, or 2023 and only eight for 2023. Therefore, the overview of congenital syphilis in the WHO European Region is not clear due to the low data availability among non-EU/EEA countries(8).

To improve baseline data on CS cases and to understand CS surveillance in non-EU/ EEA countries, the WHO Regional Office for Europe conducted a survey on congenital syphilis in 2024.

### WHO survey on congenital syphilis in non-EU/EEA countries

The WHO Regional Office for Europe conducted a survey using “WHO Annual Reporting Form on Sexually Transmitted Infections for the period January-December 2021-2023”. The reporting form was sent through official correspondence to the 24 countries in the WHO European region that, as of June 2024, did not belong to EU/EEA: Albania, Andorra, Armenia, Azerbaijan, Belarus, Bosnia and Herzegovina, Georgia, Israel, Kazakhstan, Kyrgyzstan, Monaco, Montenegro, North Macedonia, Republic of Moldova, Russian Federation, San Marino, Serbia, Switzerland, Tajikistan, Turkey, Turkmenistan, Ukraine, United Kingdom, and Uzbekistan (in alphabetical order). National focal points were asked to complete the form in English or Russian. The survey form is available in Supplementary File 1. The survey included questions on how CS surveillance is conducted and covered: type of data collection system (universal or sentinel), data sources (public sectors only or both public and private sectors), estimated reporting coverage [(Number of reported cases/Number of actual cases) *100 (%)] (< 25%, 26-50%, 51-75%, and 76-100%), and case definition used for surveillance. The survey also included questions about key programmatic indicators for EMTCT of syphilis, including the number and percentage of pregnant women attending ANC who were tested for syphilis, who tested positive, and who received treatment among those who tested positive.

### Survey results

We received data from 21 of 24 member states, and 19 countries agreed that their data could be published. The 19 countries were Albania, Andorra, Armenia, Azerbaijan, Belarus, Georgia, Israel, Kyrgyzstan, Monaco, Montenegro, North Macedonia, Republic of Moldova, San Marino, Serbia, Switzerland, Tajikistan, Türkiye, Ukraine and Uzbekistan (in alphabetical order).

#### Surveillance systems

Fifteen of the 19 countries reported having universal surveillance for CS, two reported sentinel surveillance, and two did not provide any information (Table 1). Twelve countries reported that data came from both the public and private sectors, six reported that data were only from the public sector, and one did not provide any information. Of 15 countries that provided estimates of the coverage of their surveillance system, 13 estimated that it was between 76% and 100%, one between 51% and 75% and one less than 25%.

**Table 1.** Congenital syphilis surveillance systems in 19 non-EU/EEA member states in the WHO European Region.

| Country | Universal or Sentinel | Data source | Coverage<sup>a</sup> | Case definition<sup>b</sup> | Stillbirth inclusion |
| --- | --- | --- | --- | --- | --- |
| Albania | Universal | Both | 76-100 | Unknown | Unknown |
| Andorra | Universal | Both | 76-100 | E | Yes |
| Armenia | Universal | Both | 76-100 | E, H, L | Yes |
| Azerbaijan | Universal | Public | 76-100 | L | No |
| Belarus | Universal | Both | 76-100 | Unknown | NDR |
| Georgia | Universal | Both | 76-100 | E, H, L | No |
| Israel | Universal | Public | NDR | H, L | No |
| Kyrgyzstan | NDR | Both | 76-100 | E, H, L | Yes |
| Monaco | Sentinel | Public | <25 | Unknown | Unknown |
| Montenegro | Universal | Both | 51-75 | EU 2018 | No |
| North Macedonia | Universal | Both | NDR | EU 2018<sup>c</sup> | Unknown |
| Republic of Moldova | Universal | Both | 76-100 | H, L | Yes |
| San Marino | Universal | Public | 76-100 | Unknown | NDR |
| Serbia | Universal | Both | 76-100 | EU 2018 | No |
| Switzerland | Universal | Both | 76-100 | H, L | Yes |
| Tajikistan | NDR | NDR | NDR | Unknown | Unknown |
| Türkiye | Universal | Both | 76-100 | E,L | Unknown |
| Ukraine | Universal | Public | NDR | Unknown | No |
| Uzbekistan | Sentinel | Public | 76-100 | Global<sup>d</sup> | Yes |

NDR: no data reported
a. Countries were asked to choose an estimated coverage [(Number of reported cases/Number of actual cases) \*100 (%)] from < 25%, 26-50%, 51-75%, or 76-100%.
b. E: examination, H: clinical history, L: laboratory
c. 2018 EU case definition (see Box 1)
d. global surveillance case definition (see Box 1)

#### Case definitions

Thirteen countries provided details of their CS case definition. Three countries used the 2018 EU case definition (see Box 1), and one country reported its definition was aligned with the global surveillance case definition. Nine countries reported having their own case definition. Eight of the nine (see Table 1) reported that their definition incorporated laboratory findings, five clinical examinations, and six clinical history. Six of the 12 countries that provided information on whether or not their CS definition included stillbirths reported that stillbirths were not included.

#### Reported cases

Table 2 shows the number of CS reported by 19 countries in the 2024 survey for 2021, 2022 and 2023. Table 2 also provides country estimates of CS cases per 100,000 live births using live birth estimates from the United Nations World Population Prospects 2024 (9). Seven out of 19 countries (37%) reported that they did not have any CS cases in 2021, 2022 or 2023. The maximum reported case rate was 22.9 per 100,000 live births. The total number of CS cases reported by the 19 countries increased from 61 (1.7 per 100,000 live births) in 2021 to 93 (2.8 per 100,000 live births) in 2023.

**Table 2.** Total number of reported congenital syphilis cases and congenital syphilis cases per 100,000 live births among the non-EU/EEA member states in the WHO European Region, 2021-2023.

| Country | 2021 |  | 2022 |  | 2023 |  |
| --- | --- | --- | --- | --- | --- | --- |
|  | Number | Rate | Number | Rate | Number | Rate |
| Albania | 0 | 0.0 | 0 | 0.0 | 0 | 0.0 |
| Andorra | 0 | 0.0 | 0 | 0.0 | 0 | 0.0 |
| Armenia | 1 | 2.8 | 2 | 5.7 | 6 | 17.1 |
| Azerbaijan | 4 | 2.9 | 11 | 8.5 | 25 | 20.0 |
| Belarus | 0 | 0.0 | 0 | 0.0 | 0 | 0.0 |
| Georgia | 2 | 4.1 | 1 | 2.3 | 2 | 4.5 |
| Israel | 2 | 1.1 | 6 | 3.4 | 6 | 3.5 |
| Kyrgyzstan | 2 | 1.3 | 3 | 2.0 | 4 | 2.6 |
| Monaco | 0 | 0.0 | 0 | 0.0 | 0 | 0.0 |
| Montenegro | 0 | 0.0 | 0 | 0.0 | 0 | 0.0 |
| North Macedonia | 0 | 0.0 | 1 | 5.3 | 1 | 5.9 |
| Republic of Moldova | 8 | 22.9 | 7 | 21.9 | 2 | 6.1 |
| San Marino | 0 | 0.0 | 0 | 0.0 | 0 | 0.0 |
| Serbia | 1 | 1.6 | 0 | 0.0 | 0 | 0.0 |
| Switzerland | 1 | 1.1 | 0 | 0.0 | 1 | 1.2 |
| Tajikistan | 1 | 0.4 | 4 | 1.5 | 1 | 0.4 |
| Türkiye | 38 | 3.3 | 47 | 4.4 | 44 | 4.1 |
| Ukraine | 1 | 0.3 | 2 | 0.9 | 1 | 0.5 |
| Uzbekistan | 0 | 0.0 | 0 | 0.0 | 0 | 0.0 |
| **Total** | **61** | **1.7** | **84** | **2.5** | **93** | **2.8** |

NDR: no data reported
All data from the 2024 survey unless otherwise specified.

#### EMTCT programmatic data

Table 3 provides EMTCT programmatic data. Seven countries provided data in the 2024 survey, and three countries provided data in the GAM. Of the nine countries with syphilis testing coverage data for 2023, five reported testing coverage above the EMTCT validation target of 95%. Among the seven countries that reported treatment coverage data, six of these reported that coverage was above 95%. Overall, three countries reported both testing and treatment coverage above 95%, and two of these, the Republic of Moldova and Belarus, have been validated for EMTCT of syphilis.

**Table 3.**
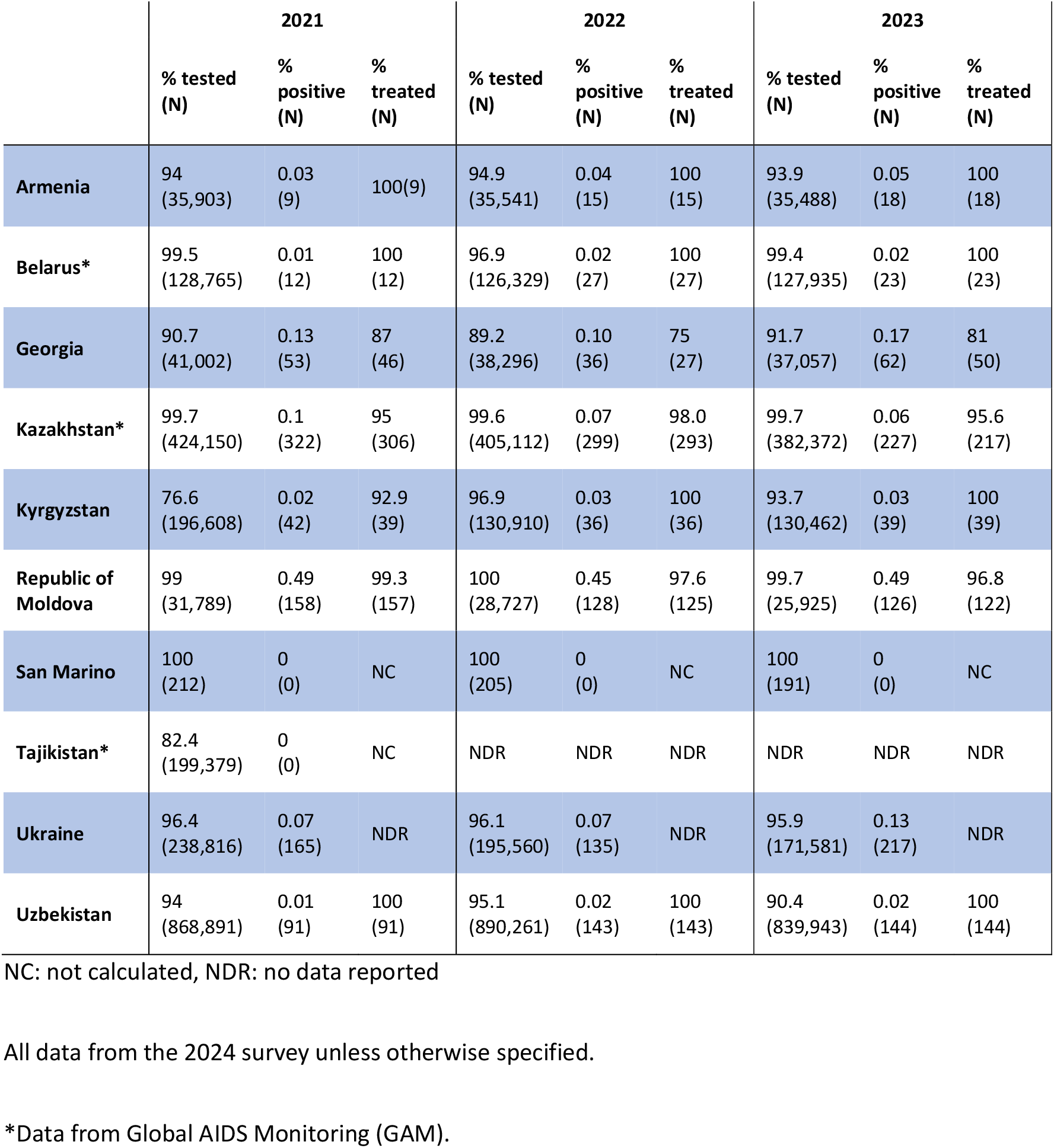
Coverage of syphilis testing, test positivity and treatment coverage in women attending antenatal care services in 10 countries with available data, 2021-2023.

#### Gaps in congenital syphilis data

Global AIDS Monitoring (GAM) annually collects congenital syphilis case numbers. Comparing country reporting status with GAM (Figure 1), this study shows improved reporting; 11 countries not reporting to GAM during 2021-2023 provided data to the WHO Regional Office for Europe (2 countries declined to publish their data). Additionally, five countries inconsistently reporting to GAM from 2021 to 2023 provided case numbers. EMTCT programmatic data, also covered by GAM, includes eight reporting countries, with three (Kyrgyzstan, San Marino, Uzbekistan) not reporting to GAM from 2021 to 2023 and providing data in this survey.

**Figure 1.**
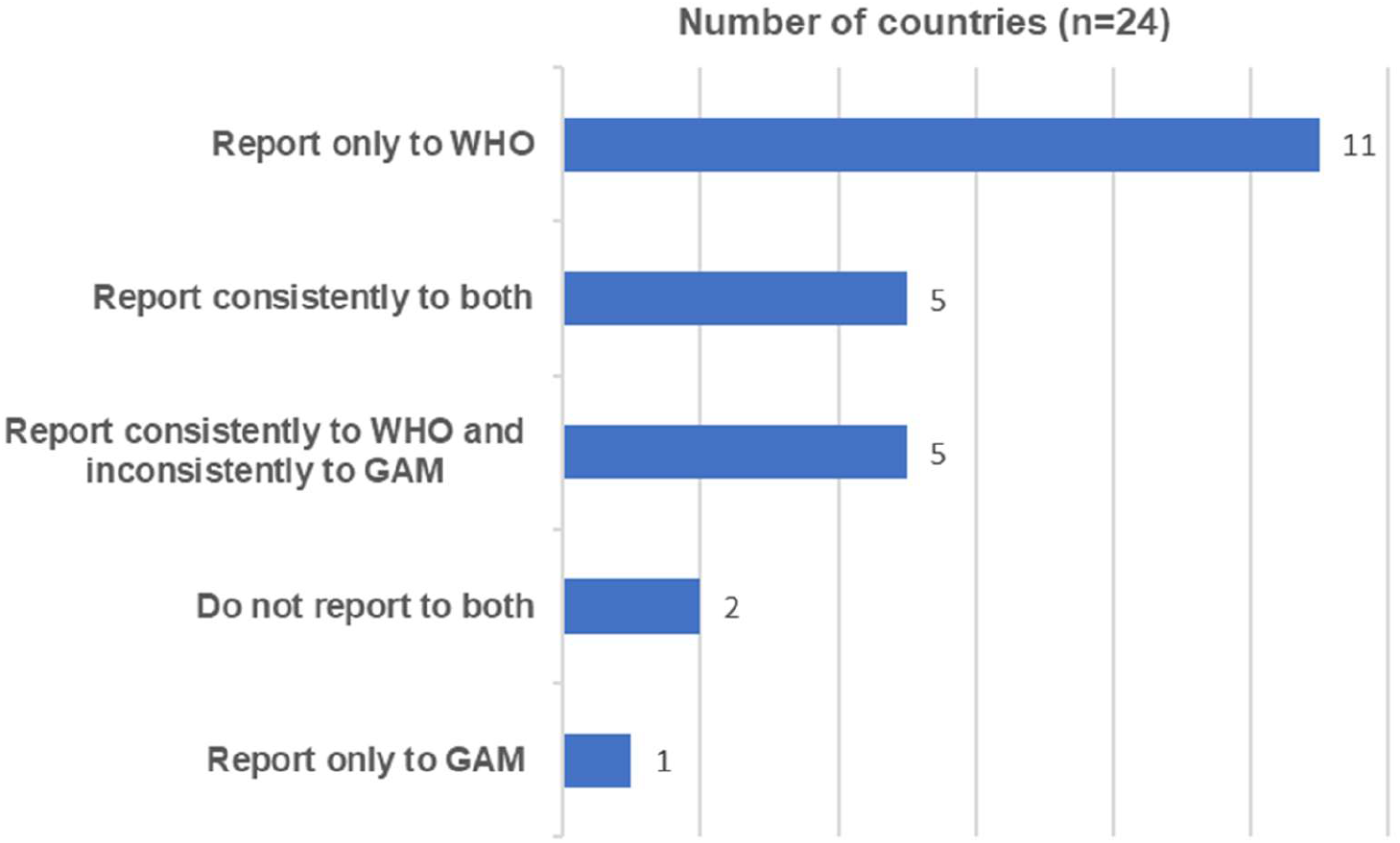
Congenital syphilis reporting status to WHO Regional Office for Europe and Global AIDS Monitoring in non-EU/EEA countries, 2021-2023. GAM: Global AIDS Monitoring. Data source: Global AIDS Monitoring in 2021,2022,2023 and WHO Annual Reporting Form on Sexually Transmitted Infections for the period January-December 2021-2023.

### Gaps in CS surveillance and implications

Various challenges in CS surveillance in the non-EU/EEA countries were identified. The comprehensiveness and coverage of CS surveillance vary between countries, and there is a need to strengthen the reporting coverage, especially from the private sector. Another issue was the difference in case definitions used for surveillance. While the most commonly used definition among EU/EEA countries is the 2018 EU case definition, which was used in 14 out of 26 reporting countries in 2023 (6), there was a huge variation in the CS case definition among non-EU/EEA countries. Having a uniform definition used across the WHO European region would benefit regional efforts to EMTCT of syphilis by enabling country comparisons.

We observed an increase in the CS notification rate in the non-EU/EEA. Although the rate remains below the global target of 50 cases per 100,000 live births by 2030, it is above the 2030 European region target of 1.0 case per 100,000 live births. It should also be noted that the actual incidence could be higher due to several contributing factors. First, the comprehensiveness and coverage of CS surveillance in the non-EU/EEA countries were suboptimal, and the whole population was not covered in many countries. Second, the case definition used for surveillance was not standardised, which could have hindered an accurate understanding of the CS epidemic in the non-EU/EEA. Moreover, while a previous study showed that substantial proportions (from 33% to 81%) of syphilis-exposed pregnancies resulted in stillbirth or neonatal death(10), only seven among 19 countries included stillbirth cases. This gap in stillbirth inclusion could also have contributed to the underestimation. Third, the coverage of syphilis testing in ANC visits was below 95% in four countries, and only 10 out of 24 countries had available coverage data for at least one year between 2021 and 2023. Therefore, some CS cases could have been missed due to a lack of access to syphilis testing during pregnancy. Strengthening CS surveillance is essential to understanding the accurate CS burden in the non-EU/EEA countries.

Data on the programmatic indicators, which are essential for assessing progress towards EMTCT, were very limited, which could be a challenge for monitoring and evaluating progress towards EMTCT of syphilis. This may be due to various reasons, including incomplete reporting at the facility or regional levels, lack of coordination between different health sectors and low priority given to national reporting on CS. Further work is needed to see if these data are readily available through the national routine health information system.

The 2024 survey results provided valuable insights into the current situation and challenges in CS surveillance in the non-EU/EEA countries of the WHO European Region. While we employed a uniform data collection method and achieved a high response rate, it is important to consider the high heterogeneity between reporting countries in various ways, including surveillance systems, CS definitions used for surveillance, access to ANC testing and treatment, testing policies, access to STI testing, diagnostic techniques and reporting practices. The observed improvement in the data availability in the number of CS cases and EMTCT programmatic data in our study suggested the importance of a coordinated international reporting system dedicated specifically to STIs.

## Conclusions

Along with a rise in the CS notification rate, various challenges in CS surveillance in the non-EU/EEA countries of the WHO European Region were identified. Enhanced commitments and coordinated approaches in strengthening the CS surveillance system at the international level are needed to ensure the region maintains the course towards achieving the elimination of EMTCT of syphilis in the region.

## Statements

### Ethical statement

Ethical approval was not necessary since we used secondary data sources.

### Use of artificial intelligence tools

None declared

### Data availability

All available data collected in this study can be shared upon reasonable request after acquiring approval from the respective countries.

## Acknowledgements

WHO European Region non-EU/EEA STI Surveillance network members:

Albania: Adela Vasili;

Andorra: Jennifer Fernández Garcia;

Armenia: Hovhannes Hovhannisyan;

Azerbaijan: Oleg Salimov;

Belarus: Hanna P. Muzychenka;

Georgia: Ketevan Galdavadze;

Israel: Rivka Rich;

Kyrgyzstan: Elvira Torobekova;

Monaco: Thomas Althaus;

Montenegro: Alma Cicic and Milena Lopicic;

North Macedonia: Dragan Kochinski;

Republic of Moldova: Aurelia Popov;

San Marino: Claudio Muccioli;

Serbia: Danijela Simic;

Switzerland: Jean-Luc Richard and Thibault Lovey;

Tajikistan: Sakina Shoeva;

Türkiye: Taliha Karakök and Mehmet Balci;

Ukraine: Liudmila Polanska;

Uzbekistan: Dilmurad Zhumanov.

The views and opinions expressed in this paper are those of the authors and not necessarily the views, decisions or policies of the World Health Organization (WHO)

## Authors’ contributions

MO led the data collection process, wrote up the manuscript, and generated tables and figures.

All other authors contributed to the data collection, reviewed the manuscript and agreed on its final version.

