## Supplementary FIle for "Towards the elimination of mother-to-child transmission of syphilis in the WHO European region: Implications from a survey in the non-European Union and European Economic Area countries"

### Supplementary File 1. WHO Annual Reporting Form on Sexually Transmitted Infections for the period January-December- 2021-2023

**1-3. Registered cases of sexually transmitted infections in 2021, 2022, 2023 (one table for each year)**

| **Disease** | **Gender** | **Female** | | | **Male** | | | | **Unknown** |
| --- | --- | --- | --- | --- | --- | --- | --- | --- | --- |
|  |  | transmission route | | | transmission route | | | | Total |
|  | By age group | Heterosexual | Other | Unknown | MSM | Heterosexual | Other | Unknown |  |
| **Syphilis**  **(Primary, Secondary, Early latent)** | 0-14 |  |  |  |  |  |  |  |  |
|  | 15-19 |  |  |  |  |  |  |  |  |
|  | 20-24 |  |  |  |  |  |  |  |  |
|  | 25-34 |  |  |  |  |  |  |  |  |
|  | 35-44 |  |  |  |  |  |  |  |  |
|  | 45+ |  |  |  |  |  |  |  |  |
|  | unknown |  |  |  |  |  |  |  |  |
| **Gonorrhoea** | 0-14 |  |  |  |  |  |  |  |  |
|  | 15-19 |  |  |  |  |  |  |  |  |
|  | 20-24 |  |  |  |  |  |  |  |  |
|  | 25-34 |  |  |  |  |  |  |  |  |
|  | 35-44 |  |  |  |  |  |  |  |  |
|  | 45+ |  |  |  |  |  |  |  |  |
|  | unknown |  |  |  |  |  |  |  |  |
| **Chlamydia (excluding Lymphogranuloma venereum)** | 0-14 |  |  |  |  |  |  |  |  |
|  | 15-19 |  |  |  |  |  |  |  |  |
|  | 20-24 |  |  |  |  |  |  |  |  |
|  | 25-34 |  |  |  |  |  |  |  |  |
|  | 35-44 |  |  |  |  |  |  |  |  |
|  | 45+ |  |  |  |  |  |  |  |  |
|  | unknown |  |  |  |  |  |  |  |  |
| **Lymphogranuloma venereum (LGV)** | 0-14 |  |  |  |  |  |  |  |  |
|  | 15-19 |  |  |  |  |  |  |  |  |
|  | 20-24 |  |  |  |  |  |  |  |  |
|  | 25-34 |  |  |  |  |  |  |  |  |
|  | 35-44 |  |  |  |  |  |  |  |  |
|  | 45+ |  |  |  |  |  |  |  |  |
|  | unknown |  |  |  |  |  |  |  |  |

**4. Congenital syphilis**

**4.1. Please indicate the total number of congenital syphilis cases reported in 2021-2023 and whether stillbirths are included or not.**

| **Year** | **The total number of cases** | **Stillbirths included?** |
| --- | --- | --- |
| **2021** |  | **( Yes No Unknown )** |
| **2022** |  | **( Yes No Unknown )** |
| **2023** |  | **( Yes No Unknown )** |

**4.2. Please indicate the case definition of congenital syphilis if it is specified in reporting.**

**4.3. Please indicate the percentage and number of pregnant women attending antenatal care who were screened for syphilis, the percentage that tested positive, and the percentage that was treated.**

| **Year** | **Number tested** | **Percentage tested** | **Number positive** | **Percentage positive** | **Number treated** | **Percentage treated** |
| --- | --- | --- | --- | --- | --- | --- |
| **2021** |  | **%** |  | **%** |  | **%** |
| **2022** |  | **%** |  | **%** |  | **%** |
| **2023** |  | **%** |  | **%** |  | **%** |

**5. Priority populations**

**5.1 Men who have sex with men**

Is there any ongoing or completed study in the last five years on the prevalence and/or trend of **syphilis** or **gonorrhoea** among **men who have sex with men**? If so, please provide information and key results.

**5.2 Female sex workers**

Is there any ongoing or completed study in the last five years on the prevalence and/or trend of **syphilis** or **gonorrhoea** among **female sex workers**? Please indicate the data and source, if any.

**5.3 Other populations**

Are there any studies on the prevalence and/or trend of syphilis or gonorrhoea among **other specific populations** in the last five years? If so, please provide details on the population and results.

**6. Surveillance system**

**6.1. Please fill in the table below about the surveillance system and data source.**

**Please note that universal reporting implies that all healthcare providers who diagnose STIs are expected to report, while sentinel reporting means that reporting is done only by selected healthcare facilities.*

| ***Disease*** | ***Surveillance system*** | ***Data source*** |
| --- | --- | --- |
| Syphilis  (Primary, Secondary, Early latent) | (universal sentinel) | (Only public sector, Public and private sector) |
| Gonorrhoea | (universal sentinel) | (Only public sector, Public and private sector) |
| Chlamydia  (excluding Lymphogranuloma venereum (LGV)) | (universal sentinel) | (Only public sector, Public and private sector) |
| Lymphogranuloma venereum (LGV) | (universal sentinel) | (Only public sector, Public and private sector) |
| Congenital syphilis | (universal sentinel) | (Only public sector, Public and private sector) |

**6.2. Please choose the estimated reporting coverage* of each disease from below 25%, 26-50%, 51-75%, and 76-100%.**

**(Number of reported cases/Number of actual cases) *100 (%)*

| **Disease** | **Estimated coverage (%)** |
| --- | --- |
| Syphilis (Primary, Secondary, Early latent) | <25, 26-50, 51-75, 76-100 |
| Gonorrhoea | <25, 26-50, 51-75, 76-100 |
| Chlamydia (excluding Lymphogranuloma venereum (LGV)) | <25, 26-50, 51-75, 76-100 |
| Lymphogranuloma venereum (LGV) | <25, 26-50, 51-75, 76-100 |
| Congenital syphilis | <25, 26-50, 51-75, 76-100 |

**6.3.** **Is your country reporting Neisseria gonorrhoea antimicrobial resistance (AMR) to the WHO Gonococcal Antimicrobial Surveillance Programme (GASP)? If not, please describe the challenges to participating.**

**6.4. Is there any surveillance system for AMR surveillance for Neisseria gonorrhoeae in your country? If so, please provide details.**

**Any additional comments**
